# Reduced neutralisation of SARS-COV-2 Omicron-B.1.1.529 variant by post-immunisation serum

**DOI:** 10.1101/2021.12.10.21267534

**Authors:** Wanwisa Dejnirattisai, Robert H Shaw, Piyada Supasa, Chang Liu, Arabella SV Stuart, Andrew J Pollard, Xinxue Liu, Teresa Lambe, Derrick Crook, Dave I Stuart, Juthathip Mongkolsapaya, Jonathan S Nguyen-Van-Tam, Matthew D Snape, Gavin R Screaton, the Com-COV2 study group

**Author notes:** contributed equally and are joint first authors (Wanwisa Dejnirattisai, Robert H Shaw, Piyada Supasa). joint senior author (Matthew D Snape, Gavin R Screaton).

## Abstract

In this report, we present live neutralisation titres against SARS-CoV-2 Omicron variant, compared with neutralisation against Victoria, Beta and Delta variants. Sera from day-28 post second-dose were obtained from participants in the Com-COV2 study who had received a two-dose COVID-19 vaccination schedule with either AstraZeneca (AZD1222) or Pfizer (BNT162b2) vaccines. There was a substantial fall in neutralisation titres in recipients of both AZD1222 and BNT16b2 primary courses, with evidence of some recipients failing to neutralise at all. This will likely lead to increased breakthrough infections in previously infected or double vaccinated individuals, which could drive a further wave of infection, although there is currently no evidence of increased potential to cause severe disease, hospitalization or death.

---

SARS-CoV-2 is estimated to have caused 265 million infections and 5.25 million deaths over the last 2 years^1^. Following the sharing of the sequence of the original SARS-CoV2 strain in January 2020, an unprecedented effort led to the production of vaccines, seven of which are currently approved by the WHO for emergency use with an estimated 8.18 billion doses delivered. All of the current vaccines are based around the original SARS-CoV2 strain and are designed primarily to raise an antibody response against the spike protein (S), although T-cell responses are also elicited and may play a role in protection from severe disease.

Since the SARS-CoV-2 RNA polymerase is intrinsically error prone mutation of the viral genome is to be expected. Changes that lead to increased viral fitness conferring, for example, increased transmissibility or the ability to evade the host immune response, undergo rapid natural selection. Over the last 2 years, genomic surveillance has uncovered thousands of individual mutations of the SARS-CoV-2 genome at multiple locations. Mutations in S are of particular concern, as they may lead to increased transmission or immune escape. Towards the end of 2020, viral variants containing multiple changes in S were reported: Alpha (first detected in UK), Beta (South Africa), Gamma (Brazil) and Delta (India). These variants contain mutations in the Receptor Binding Motif (RBM), a small 25 amino acid patch at the tip of S that mediates interaction with the ACE2 receptor (α−1, β−3, γ−3, δ −2). These changes may lead to increased transmissibility by increasing affinity to ACE2 (α−7x, β−19x, γ−19x, δ−2x)^2^ or lead to immune escape. First Alpha then and then Delta spread worldwide and fuelled successive waves of infection, whilst Beta and Gamma led to large localised outbreaks in Southern Africa and South America respectively, but did not dominate elsewhere.

Currently, Delta is estimated to represent over 99% of infections worldwide, however, a new variant of concern Omicron(B.1.1.529) was first reported from South Africa on 24 November 2021^3^, but has since been reported in multiple countries and is causing great concern. Early reports from South Africa suggest Omicron is highly transmissible, in a population where 60-80% already show serological evidence of previous infection or vaccination, and it is clear that Omicron is able to break through natural and vaccine-induced immunity and vaccines to some extent, although early reports are not pointing to more severe disease.

Omicron contains a large number of mutations in S compared to previous variants of concern, 30 amino acid substitutions, deletion of 6 residues and insertion of 3 residues^2^. Mutations are concentrated around the RBM where they are predicted to increase affinity for ACE2 and lead to reduced neutralisation activity of RBM-binding antibodies which block interaction with ACE2. Mutations are also present at other sites for binding of potent neutralising antibodies in the Receptor Binding Domain and N-Terminal Domain. The large mutation burden in Omicron S, suggest mutations have evolved to increase ACE2 affinity and evade the antibody response. There is concern that Omicron will lead to increased propensity to infect individuals, who have received vaccines based on the original S sequence.

In this report we have performed neutralisation assays using an isolate of Omicron obtained from an infected case in the UK. Neutralisation assays were performed on sera from seronegative individuals (defined by anti-nucleocapsid IgG) vaccinated with two doses of the Oxford-AstraZeneca vaccine AZD1222 (n=22), or Pfizer-BioNTech BNT162b2 vaccine (n=21) Samples were obtained four weeks following the second dose of vaccine administered 8-11 (median 9.0) weeks after the first as part of the Com-COV2 study (Table S1)^4^

Live virus neutralisation titres against Omicron are compared with neutralisation against Victoria, an early pandemic SARS-CoV-2 strain together with neutralization against Beta and Delta variants. Neutralising titres on sera from participants who had received homologous AZD1222 dropped to below the detectable threshold in all but one participant (Figure 1A, B). Median neutralising titres on sera from participants who had received homologous BNT162b2 dropped 29.8-fold from 1609 (Victoria strain) to 54 (Omicron variant), with one participant dropping below the detection threshold. In most cases in the samples that failed to neutralize with FRNT50 <1/20 some residual neutralizing activity was detected (Figure 1C)

**Figure 1.**
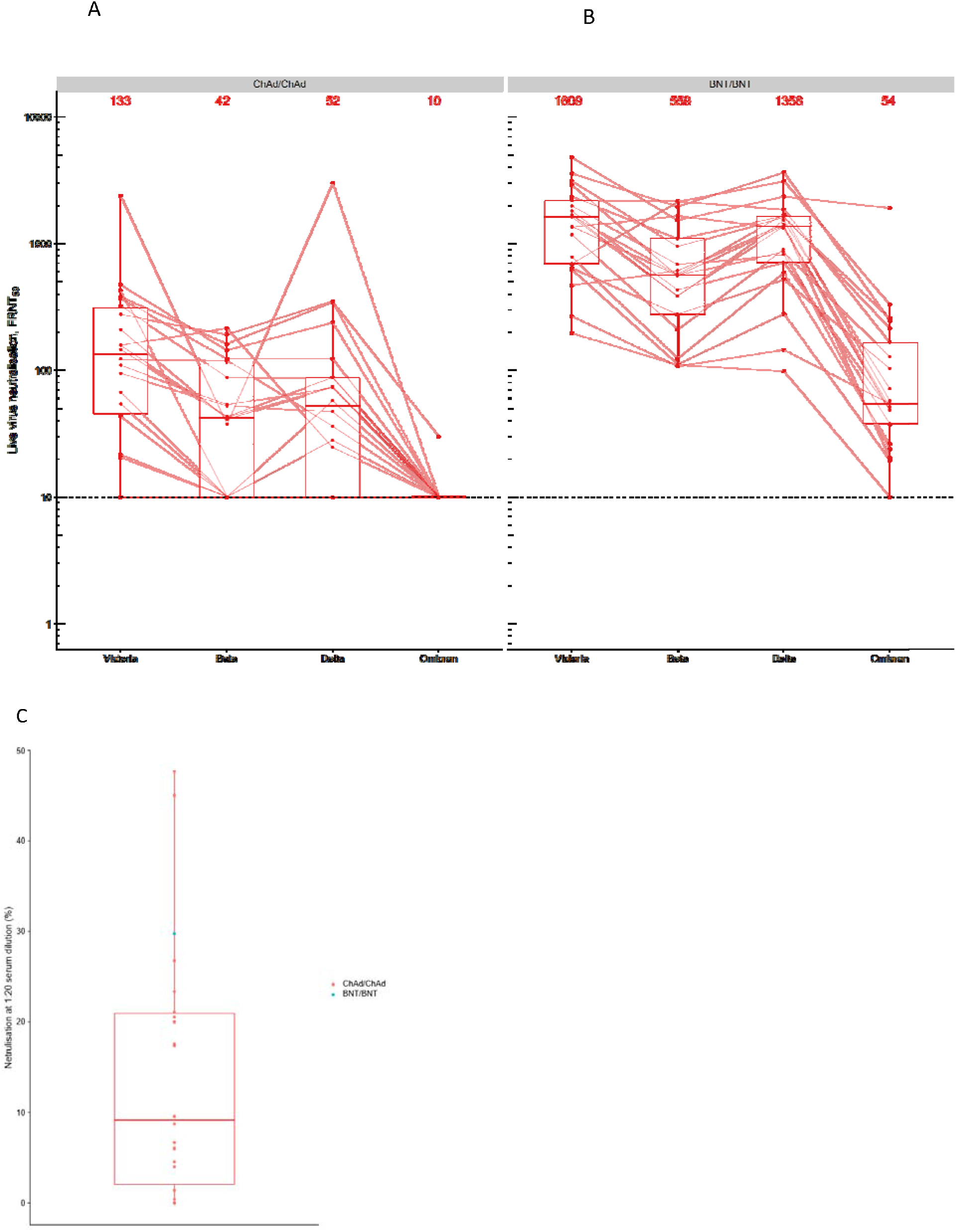
Neutralisation assays of SARS-CoV-2 Omicron. Neutralisation of Victoria, Beta, Delta and Omicron using (A) AZD1222 serum (ChAd) and (B) BNT162b2 (BNT) serum. Median values are indicated above each column. The data underpinning the Victoria, Beta and Delta neutralisation have been previously reported^4^. The horizontal dotted line indicates half the value of the lower limit of detection. (C) Percent neutralization at serum dilution of 1/20 for those sera which failed to achieve FRNT50 at 1/20, the green dot represents the single BNT162b2 sample.

In summary, there was a substantial fall in neutralisation titres in recipients of both homologous AZD1222 and BNT162b2 primary courses, with evidence of some recipients failing to neutralise at all. These data suggest Omicron is more antigenically distant from the original SARS-CoV2 vaccine strain than the previously most distant strains Beta and Delta, and are consistent with recently published data for recipients of BNT162b2^5^. This will likely lead to increased breakthrough infections in previously infected or double vaccinated individuals, which could drive a further wave of infection, although there is currently no evidence of increased potential to cause severe disease, hospitalization or death. The impact on disease severity is currently unknown but other aspects of the immune system such as non-neutralising antibodies and cellular immunity are not expected to be as severely impacted and may confer a degree of protection against hospitalisation and severe disease. However, it should be noted that higher transmission, will inevitably lead to increased numbers of cases and greater burden on health systems, even without proportional changes in severity. The impact of a third-dose vaccine is currently unknown but may be expected to raise neutralization titres against the new variant; testing of samples from Cov-Boost^6^ to provide these data is ongoing and will provide further understanding of the potential for a boosting strategy as a control measure for Omicron infection and transmission.

The immune escape reported here may lead Omicron to displace Delta to become the dominant strain worldwide. If this were to occur it may be necessary to produce vaccines tailored to Omicron, however, because of the antigenic distance of Omicron, these might be unlikely to give protection against previous strains. This may stimulate consideration of a switch from the current monovalent vaccine strategy towards multivalent formulations currently used in seasonal influenza vaccines. Finally, from the data presented here, it is clear that possessing a high starting titre against early pandemic strains gives a higher level of neutralisation of Omicron, which could be obtained by deploying third booster doses of vaccine. Reaching the unvaccinated with current vaccines remains a priority in order to reduce transmission levels and reduce the potential for severe disease in the immunologically naïve.

## Data Availability

Upon requests directed to the corresponding author, after approval of a proposal, data can be shared through a secure online platform.

