## Supplementary appendix for "Reduced neutralisation of SARS-COV-2 Omicron-B.1.1.529 variant by post-immunisation serum"

**Supplementary file**

Table S1 Baseline characteristics by vaccine schedules

|  | AZD1222/AZD1222  N=22 | BNT162b2/BNT162b2  N=21 |
| --- | --- | --- |
| Interval between 1^st^ and 2^nd^ dose, Median(range), weeks | 9.5 (8-11) | 9 (8-10) |
| Age, Median(range), years | 61.9 (51.6-69.2) | 60.8 (51.2-69.5) |
| Sex, female | 10 (45.4%) | 7 (33.3%) |
| Ethnicity, white | 17 (77.2%) | 19 (90.5%) |

**Methods**

*The Com-COV2 study*

Com-COV2 (ISRCTN:27841311) is an ongoing, multi-centre, single-blinded UK study enrolling individuals aged 50 years or older previously immunized with a single dose of AZD1222 or BNT162b2 at least 8 weeks previously. Participants were randomized on a 1:1:1 basis to receive the same dose again, or a COVID-19 vaccine manufactured by Novavax or Moderna (Ethics reference 21/SC/0119). Samples were selected for the AZD1222/AZD1222 and BNT162b2/BNT162b2 groups from an immunology cohort (25 per group), pre-defined at the point of enrolment.

*Viral culture and isolation*

Omicron virus was cultured from a routine diagnostic throat swab (IRAS Project ID: 269573, Ethics Ref: 19/NW/0730. Briefly, VeroE6/TMPRSS2 cells (NIBSC) were maintained in Dulbecco's Modified Eagle Medium (DMEM) high glucose supplemented with 1% fetal bovine serum, 2mM Glutamax, 100 IU/ml penicillin-streptomycin and 2.5ug/ml amphotericin B, at 37 °C in the presence of 5% CO2 before inoculation with 200ul of swab fluid. Cells were further maintained at 37°C with daily observations for cytopathic effect (CPE). Virus containing supernatant were clarified at 80% CPE by centrifugation at 3,000 r.p.m. at 4 °C before being stored at -80 °C in single-use aliquots. Viral titres were determined by a focus-forming assay on Vero CCL-81 cells (ATCC). Sequencing of the Omicron isolate shows the expected consensus S gene changes (A67V, Δ69-70, T95I, G142D/Δ143-145, Δ211/L212I, ins214EPE, G339D, S371L, S373P, S375F, K417N, N440K, G446S, S477N, T478K, E484A, Q493R, G496S, Q498R, N501Y, Y505H, T547K, D614G, H655Y, N679K, P681H, N764K, D796Y, N856K, Q954H, N969K, L981F), an intact furin cleavage site and a single additional mutation A701V

*Focus reduction neutralisation test*

The focus reduction neutralisation test (FRNT) was carried out in order to determine the neutralisation potency of tested sera using passage 1 virus with the sequence above. Tissue culture 96-well, flat-bottom microplates were seeded with Vero CCL-81 cells 24 h prior to use and were used at around 90-95% confluent monolayer. A three-fold dilution of heat-inactivated serum samples, starting from 1:20, were pre-mixed with an equal volume of viral solution containing 100 foci. The controls included virus alone layered on top of Vero CCL-81cells. After 1 hour of incubation at 37°C, the serum-virus mixture at each dilution was added in duplicate to Vero cells monolayer and incubated for 2 hrs followed by the addition of 1.5% semi-solid carboxymethyl cellulose (CMC) immobilizing medium to each well to prevent virus spread. After addition of the overlay, cells were further incubated for 20 hrs.

The viral foci were visualized by focus-forming assay. Briefly, cells were fixed and permeabilized with 4% PFA and 2% triton-X 100, respectively. This was followed by the staining with human anti-N mAb (mAb206) and peroxidase-conjugated goat anti-human IgG (A0170; Sigma). Finally, TrueBlue Peroxidase substrate was used to develop the plates followed by imaging and counting the foci under ELISPOT reader. The FRNT50 titer is calculated using the probit program from the SPSS package.

*Acknowledgements:*

The work was supported by the Chinese Academy of Medical Sciences (CAMS) Innovation Fund for Medical Science (CIFMS), China (grant number: 2018-I2M-2-002) to DIS and GRS, DIS by the UKRI MRC (MR/N00065X/1). DIS and GRS are Jenner Investigators. The Wellcome Centre for Human Genetics is supported by the Wellcome Trust (grant 090532/Z/09/Z). Com-COV2 was funded by the UK Government through the National Institute for Health Research (NIHR), the Vaccine Task Force (VTF) and the Coalition for Epidemic Preparedness Innovations (CEPI). This research was supported by the NIHR Oxford Biomedical Research Centre, NIHR Guy's and St Thomas' Biomedical Research Centre, NIHR King's Clinical Research Facility and NIHR Policy Research Programme (PR-R17-0916-22001), the Southampton NIHR Biomedical Research Centre and Southampton NIHR Clinical Research Facility and delivered through the NIHR-funded National Immunisation Schedule Evaluation Consortium (NISEC). NVX-CoV2373 vaccine was supplied for use in Com-COV2 by Novavax, Inc. MDS and AJP are Jenner Investigators and NIHR Senior Investigators and supported by the Oxford NIHR Biomedical Research Centre.

*Declarations of interest:*

G.R.S sits on the GSK Vaccines Scientific Advisory Board and is a founder member of RQ Biotechnology. MDS acts on behalf of the University of Oxford as an investigator on studies funded or sponsored by vaccine manufacturers, including AstraZeneca, GlaxoSmithKline, Pfizer, Novavax, Janssen, Medimmune, and MCM Vaccines. He receives no personal financial payment for this work. AJP reports grants from UKRI, CEPI and NIHR, during the conduct of the study. AJP is Chair of DHSC’s Joint Committee on Vaccination & Immunisation (JCVI), but does not participate in discussions on COVID19 vaccines, and is a member of the WHO’s SAGE. The views expressed in this article are those of the authors and do not necessarily represent the views of DHSC, JCVI, NIHR or WHO. The University of Oxford has entered into a partnership with AstraZeneca for the development of a coronavirus vaccine. JSN-V-T is seconded to the Department of Health and Social Care, England (DHSC). The views expressed in this manuscript are those of its authors and not necessarily those of DHSC.

*Contribution*

GRS and JM designed the experiment, which was performed by WD, PS and CL. WD and PS isolated the Omicron strain. DC sequenced the Omicron virus. WD, PS and CL performed and analysed the neutralising experiments. MDS is the chief investigator for Com-COV2. MDS, JVT, ASVS, RHS, TL and XL contributed to the protocol and design of Com-COV2. ASVS, RHS and RW led the implementation of Com-COV2. XL did the statistical analysis and has verified the underlying data. GRS, JM, AJP, XL and MDS drafted the manuscript, with contributions from DIS. All authors reviewed and approved the final manuscript. All authors had full access to all the data in the study and had final responsibility for the decision to submit for publication.

| **Com-COV2 Study Group** | | |
| --- | --- | --- |
| Sarah D. | Birch | Department of Infection and Tropical Medicine, Sheffield Teaching Hospitals NHS Foundation Trust |
| Anna L | Goodman | Department of Infection, Guy's and St Thomas' NHS Foundation Trust; MRC Clinical Trials Unit, University College London; NIHR BRC, Guy's & St Thomas' NHS Foundation Trust |
| Linda J. | Kay | Department of Infection, Immunity and Cardiovascular Disease, University of Sheffield |
| Ruth | Payne | Department of Infection, Immunity and Cardiovascular Disease, University of Sheffield, Sheffield, UK; Department of Infection and Tropical Medicine, Sheffield Teaching Hospitals NHS Foundation Trust, Sheffield, UK |
| Tom | Darton | Department of Infection, Immunity and Cardiovascular Disease, University of Sheffield; Department of Infection and Tropical Medicine, Sheffield Teaching Hospitals NHS Foundation Trust |
| Robert C. | Read | Faculty of Medicine and Institute for Life Sciences, University of Southampton; NIHR Southampton Biomedical Research Centre, University Hospital Southampton |
| Mary | Ramsay | Immunisation and Countermeasures Division, National Infection Service, UK Health Security Agency, London, UK |
| Katie J. | Drury | Infection Research Group, Hull University Teaching Hospitals NHS Trust, Hull, UK |
| Nick | Easom | Infection Research Group, Hull University Teaching Hospitals NHS Trust, Hull, UK |
| Patrick J | Lillie | Infection Research Group, Hull University Teaching Hospitals NHS Trust, Hull, UK |
| Rosa Maeve | McGing | Infection Research Group, Hull University Teaching Hospitals NHS Trust, Hull, UK |
| Tanaraj | Perinpanathan | Infection Research Group, Hull University Teaching Hospitals NHS Trust, Hull, UK |
| Annette D. | Samson | Infection Research Group, Hull University Teaching Hospitals NHS Trust, Hull, UK |
| Gemma L. | Walker | Infection Research Group, Hull University Teaching Hospitals NHS Trust, Hull, UK |
| Daniela M | Ferreira | Liverpool School of Tropical Medicine, Liverpool, UK |
| Helen | Hill | Liverpool School of Tropical Medicine, Liverpool, UK |
| Annabel | Murphy | Liverpool School of Tropical Medicine, Liverpool, UK |
| Angelina | Peterson | Liverpool School of Tropical Medicine, Liverpool, UK |
| Farah | Shiham | Liverpool School of Tropical Medicine, Liverpool, UK |
| Carla | Solórzano | Liverpool School of Tropical Medicine, Liverpool, UK |
| Jonathan | Stamen | Liverpool School of Tropical Medicine, Liverpool, UK |
| Violet | Swain | Liverpool School of Tropical Medicine, Liverpool, UK |
| Andrea M | Collins | Liverpool School of Tropical Medicine, Liverpool, UK; Liverpool University Hospitals Foundation Trust, UK |
| Paul J | Turner | National Heart & Lung Institute, Imperial College London, London, UK |
| Movin | Abeywickrema | NIHR BRC at Guy's and St Thomas' NHS Foundation Trust, London, UK |
| Alison | Davies | NIHR BRC at Guy's and St Thomas' NHS Foundation Trust, London, UK |
| Elisa | Nanino | NIHR BRC at Guy's and St Thomas' NHS Foundation Trust, London, UK |
| Alice | Packham | NIHR BRC at Guy's and St Thomas' NHS Foundation Trust, London, UK |
| Cherry | Paice | NIHR BRC at Guy's and St Thomas' NHS Foundation Trust, London, UK |
| Jo | Salkeld | NIHR BRC at Guy's and St Thomas' NHS Foundation Trust, London, UK |
| Sonia | Serrano | NIHR BRC at Guy's and St Thomas' NHS Foundation Trust, London, UK |
| Niamh | Spencer | NIHR BRC at Guy's and St Thomas' NHS Foundation Trust, London, UK |
| Jaimie | Wilson Goldsmith | NIHR BRC at Guy's and St Thomas' NHS Foundation Trust, London, UK |
| Andrea | Mazzella | NIHR BRC at Guy's and St Thomas' NHS Foundation Trust, London, UK; St George's University of London (Infection and Immunity Research Institute), London, UK |
| Saul N | Faust | NIHR Southampton Clinical Research Facility and Biomedical Research Centre, University Hospital Southampton NHS Foundation Trust, Southampton, UK; Faculty of Medicine and Institute for Life Sciences, University of Southampton, Southampton, UK |
| Robert C | Read | NIHR Southampton Clinical Research Facility and Biomedical Research Centre, University Hospital Southampton NHS Foundation Trust, Southampton, UK; Faculty of Medicine and Institute for Life Sciences, University of Southampton, Southampton, UK |
| Kirsty | Adams | NIHR UCLH Clinical Research Facility and NIHR UCLH Biomedical Research Centre, London, UK |
| Shama | Hamal | NIHR UCLH Clinical Research Facility and NIHR UCLH Biomedical Research Centre, London, UK |
| Tommy | Rampling | NIHR UCLH Clinical Research Facility and NIHR UCLH Biomedical Research Centre, London, UK |
| Marivic | Ricamara | NIHR UCLH Clinical Research Facility and NIHR UCLH Biomedical Research Centre, London, UK |
| Nicola | Turner | NIHR UCLH Clinical Research Facility and NIHR UCLH Biomedical Research Centre, London, UK |
| Vincenzo | Libri | NIHR UCLH Clinical Research Facility and NIHR UCLH Biomedical Research Centre, University College London Hospitals NHS Foundation Trust, London, UK |
| Claire H. | Brown | NIHR/Wellcome Trust Clinical Research Facility, University Hospitals Birmingham NHS Foundation Trust |
| Amisha | Desai | NIHR/Wellcome Trust Clinical Research Facility, University Hospitals Birmingham NHS Foundation Trust |
| Karishma | Gokani | NIHR/Wellcome Trust Clinical Research Facility, University Hospitals Birmingham NHS Foundation Trust |
| Kush | Naker | NIHR/Wellcome Trust Clinical Research Facility, University Hospitals Birmingham NHS Foundation Trust |
| Ehsaan | Qureshi | NIHR/Wellcome Trust Clinical Research Facility, University Hospitals Birmingham NHS Foundation Trust |
| Christopher A | Green | NIHR/Wellcome Trust Clinical Research Facility, University Hospitals Birmingham NHS Foundation Trust, Birmingham, UK; Institute of Microbiology & Infection, University of Birmingham, UK |
| Rajeka | Lazarus | North Bristol NHS Trust, Bristol, UK |
| Florentina D. | Penciu | North Bristol NHS Trust, Bristol, UK |
| Tawassal | Riaz | North Bristol NHS Trust, Bristol, UK |
| Parvinder K | Aley | Oxford Vaccine Group, Department of Paediatrics, University of Oxford, Oxford, UK |
| Elizabeth A | Clutterbuck | Oxford Vaccine Group, Department of Paediatrics, University of Oxford, Oxford, UK |
| Tanya | Dinesh | Oxford Vaccine Group, Department of Paediatrics, University of Oxford, Oxford, UK |
| Mujtaba | Ghulam Farooq | Oxford Vaccine Group, Department of Paediatrics, University of Oxford, Oxford, UK |
| Melanie | Greenland | Oxford Vaccine Group, Department of Paediatrics, University of Oxford, Oxford, UK |
| Bryn M | Horsington | Oxford Vaccine Group, Department of Paediatrics, University of Oxford, Oxford, UK |
| Yama F | Mujadidi | Oxford Vaccine Group, Department of Paediatrics, University of Oxford, Oxford, UK |
| Emma L | Plested | Oxford Vaccine Group, Department of Paediatrics, University of Oxford, Oxford, UK |
| Samuel | Provstgaard-Morys | Oxford Vaccine Group, Department of Paediatrics, University of Oxford, Oxford, UK |
| Hannah | Robinson | Oxford Vaccine Group, Department of Paediatrics, University of Oxford, Oxford, UK |
| Nisha | Singh | Oxford Vaccine Group, Department of Paediatrics, University of Oxford, Oxford, UK |
| Hanane | Trari Belhadef | Oxford Vaccine Group, Department of Paediatrics, University of Oxford, Oxford, UK |
| Iason | Vichos | Oxford Vaccine Group, Department of Paediatrics, University of Oxford, Oxford, UK |
| Rachel | White | Oxford Vaccine Group, Department of Paediatrics, University of Oxford, Oxford, UK |
| Maheshi N | Ramasamy | Oxford Vaccine Group, Department of Paediatrics, University of Oxford, Oxford, UK; Oxford University Hospitals NHS Foundation Trust, Oxford, UK |
| Natalie | Palmer | Pharmacy Clinical Trials (Adult) at Guy's and St Thomas' NHS Foundation Trust |
| J Claire | Cameron | Public Health Scotland, Glasgow, Scotland, UK |
| Neil J. | Oldfield | School of Life Sciences, University of Nottingham |
| Adam | Finn | Schools of Population Health Sciences and Cellular and Molecular Medicine, University of Bristol, Bristol, UK |
| Joann | Barker | Sheffield Teaching Hospitals NHS Foundation Trust, Sheffield, UK |
| Hayley | Colton | Sheffield Teaching Hospitals NHS Foundation Trust, Sheffield, UK |
| Jennifer N. | Hall | Sheffield Teaching Hospitals NHS Foundation Trust, Sheffield, UK |
| Kim B. | Ryalls | Sheffield Teaching Hospitals NHS Foundation Trust, Sheffield, UK |
| Nick J | Andrews | Statistics, Modelling and Economics Department, UK Health Security Agency, London, UK; Immunisation and Countermeasures Division, National Infection Service, UK Health Security Agency, London, UK |
| Maria | Allen | The Newcastle upon Tyne Hospitals NHS Foundation Trust, Newcastle upon Tyne, UK |
| Gillian | Curry | The Newcastle upon Tyne Hospitals NHS Foundation Trust, Newcastle upon Tyne, UK |
| Jeremy | Nell | The Newcastle upon Tyne Hospitals NHS Foundation Trust, Newcastle upon Tyne, UK |
| David A. | Price | The Newcastle upon Tyne Hospitals NHS Foundation Trust, Newcastle upon Tyne, UK |
| Amada | Sanchez-Gonzalez | The Newcastle upon Tyne Hospitals NHS Foundation Trust, Newcastle upon Tyne, UK |
| Hayley | Wardle | The Newcastle upon Tyne Hospitals NHS Foundation Trust, Newcastle upon Tyne, UK |
| Christopher JA | Duncan | The Newcastle upon Tyne Hospitals NHS Foundation Trust; Translational and Clinical Research Institute, Newcastle University |
| Sue | Belton | The University of Nottingham Health Service, University of Nottingham, Nottingham, UK |
| Daniel | Hammersley | The University of Nottingham Health Service, University of Nottingham, Nottingham, UK |
| Simon | Royal | The University of Nottingham Health Service, University of Nottingham, Nottingham, UK |
| David PJ | Turner | The University of Nottingham, Nottingham, UK; Nottingham University Hospitals NHS Trust, Nottingham, UK |
| Emily | Beales | The Vaccine Institute, St. George's University of London, London, UK |
| Olivia | Bird | The Vaccine Institute, St. George's University of London, London, UK |
| Eva P. | Galiza | The Vaccine Institute, St. George's University of London, London, UK |
| Paul T | Heath | The Vaccine Institute, St. George's University of London, London, UK |
| Cecilia | Hultin | The Vaccine Institute, St. George's University of London, London, UK |
| Alberto | San Francisco Ramos | The Vaccine Institute, St. George's University of London, London, UK |
| Sue | Charlton | UK Health Security Agency, Porton Down, Salisbury, UK |
| Anna | England | UK Health Security Agency, Porton Down, Salisbury, UK |
| Bassam | Hallis | UK Health Security Agency, Porton Down, Salisbury, UK |
